# Phenotypic presentation of family members of ATTRv probands and subsequent disease penetrance

**DOI:** 10.1101/2024.09.06.24313219

**Authors:** Luca Fazzini, Matteo Castrichini, Yan Li, Jose De Melo, Marta Figueiral, Jenny J. Cao, Eric W. Klee, Christian Cadeddu Dessalvi, Martha Grogan, Angela Dispenzieri, Naveen L. Pereira

## Abstract

**BACKGROUND:** Hereditary transthyretin amyloid cardiomyopathy (ATTRv-CM) is being increasingly diagnosed due to enhanced awareness and availability of newer therapeutics. Multiple *TTR* variants have been described worldwide, but with uncertain disease penetrance. The characteristics and outcomes of “previously undiagnosed” pathogenic-likely pathogenic (P/LP) *TTR* variant (genotype or G+; cardiac phenotype or P-) carriers are unknown which has important prognostic and therapeutic implications, especially for affected family members. This descriptive study aimed to delineate phenotype and cardiac penetrance in “previously undiagnosed” G+P-family members of ATTRv probands.

**METHODS:** Demographic, electrocardiographic (ECG), genetic, and imaging (echocardiography, cardiac technetium-99m pyrophosphate (PYP) and magnetic resonance imaging) data were analyzed. The prediction effect of selected baseline characteristics for ATTRv-CM development was evaluated. Kaplan-Meier and Cox regression methods were used to describe risk and predictors of ATTRv-CM development in family members.

**RESULTS:** There were 85 G+P-family members identified. Mean age was 48.5±11.7 years, 39% were male, 18% had a diagnosis of peripheral neuropathy, 15% with a history of carpal tunnel syndrome, and 4% had atrioventricular block at baseline. Of these, 55 patients had follow-up imaging studies. After a median 6.8-year follow-up, 22% developed ATTR-CM with a 10-year estimated risk of 29.5% (95% CI 7.9-46.0). Cardiac penetrance increased with increasing family member’s age. Proband’s diagnosis age (p=0.0096) and artificial intelligence (AI)-ECG prediction (p=0.0091) were promising baseline predictors of time to ATTRv-CM development.

**CONCLUSION:** In previously undiagnosed G+P-ATTRv family members, the incidence of subsequent CM is high. Predictors for CM development such as proband’s diagnosis age and AI-determined ECG probability of ATTR-CM require further investigation.

## Introduction

Cardiac amyloidosis (CA) is a well-known cause of heart failure and life-threatening disease.^1–3^ Early diagnosis of transthyretin amyloidosis (ATTR) which is a common cause of CA has become especially relevant given recent advances in diagnosis and therapy.^4^ Cardiomyopathy (CM) due to ATTR is classified as either wild-type (ATTRwt-CM) or hereditary (ATTRv-CM).^5^ Hereditary ATTR is an autosomal-dominant disease and over 130 *TTR* variants have been identified that have ethnic and geographical specific distribution.^6–8^ Different TTR genotypes are associated with varying phenotypes.^9–11^ The correlation between genetic heterogeneity, variability in disease onset, and expression remains an active area of investigation.^12^

Genetic testing is recommended in patients diagnosed with ATTR-CM in order to distinguish between wild-type and hereditary disease.^4,13–15^ Identifying a pathogenic (P) or likely-pathogenic (LP) variant not only enables a diagnosis of ATTRv-CM but also allows for cascade testing in first-degree relatives.^16^ However, managing genotype-positive (G+, carriers of a P or LP variant) phenotype-negative (P-, individuals without evidence of structural heart disease) remains a challenge due to a lack of information on the natural course of the disease and the uncertain cardiac penetrance of these *TTR* variants. Understanding outcomes and identifying high risk individuals will inform strategies for not only monitoring but also potentially treating *TTR* genetic (G+P-) carriers. The purpose of this study was to evaluate the natural history, cardiac disease penetrance, and predictors of the development of ATTRv-CM in previously undiagnosed *TTR* G+P-individuals.

## Methods

### Study population

The institutional review board of the Mayo Clinic Foundation approved this study which was a single-center, observational, and retrospective study. Among the overall population affected by amyloidosis followed at Mayo Clinic, Rochester, previously undiagnosed family members of probands who were diagnosed with ATTRv and underwent genetic testing were identified from January 1998 to October 2023. Those with P/LP *TTR* variants without evidence of ATTR-CM at baseline screening (G+P-) were included in the study. P-individuals were defined as those with the absence of echocardiographic evidence of ATTR-CM as previously described^17^ and/or negative pyrophosphate (PYP) scintigraphy. Pertinent demographic, clinical, electrocardiographic (ECG), and imaging data closest to the date of genetic testing for these patients were extracted from the electronic medical record. Patients without cardiac imaging at baseline were excluded since the baseline cardiac phenotype could not be determined. The population was divided into two groups based on the development of CM during follow-up.

### Genetic testing

Genomic DNA obtained from submitted samples was enriched for targeted regions using a hybridization-based protocol. Sequence analysis and deletion/duplication testing of the *TTR* gene were performed. A bi-directional sequence analysis was performed to test for the presence of variants in all coding regions and intron/exon boundaries of the *TTR* gene. A P/LP variant in the *TTR* gene was defined as per the American College of Medical Genetics and Genomics (ACMG) standard criteria.^18^

### Cardiomyopathy diagnosis during follow-up

The criteria used to diagnose ATTR-CM during follow-up were as follows: 1) Standard previously used echocardiographic criteria (at least two of the following features):^17^ normal left ventricular (LV) wall thickness at baseline (≤13mm) that became abnormal during follow-up; normal filling pressures (E/è≤8) that became abnormal, normal atria size at baseline that evolved to biatrial enlargement at follow-up; and normal LV global longitudinal strain (<-18%) that subsequently became abnormal. 2) a normal cardiac magnetic resonance (cMRI) scan at baseline which during follow-up demonstrated late gadolinium enhancement and increased extracellular volume consistent with ATTR-CM 3) a negative baseline PYP scintigraphy that became positive during follow-up. Patients were considered not to have developed ATTR-CM if they had any negative follow-up imaging study.

### Artificial intelligence-enabled electrocardiography

A previously developed and validated artificial intelligence (AI) algorithm to diagnose CA was applied to the baseline ECG of the G+P-carriers.^19,20^ An AI ECG was considered positive when the probability threshold to detect patients with CA by the Youden index was 0.485 or higher.

### Statistical analysis

Descriptive statistics were used to summarize the characteristics of the study population. Continuous variables are reported as mean (standard deviation, SD) or median (interquartile range [IQR]), and categorical variables are reported as proportions. To evaluate if clinical and echocardiographic characteristics are changed between baseline and follow-up, continuous variables were tested using paired two-sided t-tests, while binary variables were tested if their proportional difference between baseline and follow-up is different from 0. Kaplan-Meier method was used to estimate time-to-event models. Patients without events were censored at the last follow-up. To study the prediction effect of selected baseline characteristics, first we fit the Firth’s penalized logistic regressions using the CA development as a dependent variable and the baseline characteristics as independent variables. Next, we aimed to study if there is a synergistic effect of those baseline variables. To retain the sample size of 55, missing values of baseline variables were imputed by median values from corresponding groups (developing or not developing CA). Next, we combine these selected baseline characteristics using Ridge logistic regression. Therefore, the predictor of this Ridge logistic model is the optimal composite score of those baseline variables. We used precision-recall (PR) curve analysis to evaluate the predicting effect of the composite score for CA development. Specifically, 11-fold cross-validation (CV) was used when calculating the area under the curve (AUC), since the score was derived from an estimated model and was needed to protect against overfitting. To improve the estimated performance of the regression model, we repeated the CV procedure 100 times. Univariable Cox proportional hazard models were performed to evaluate the risk of developing overt CA. The Hazard Ratios (HR) and the corresponding 95% Confidence Intervals (CI) were reported. The *TRR* penetrance on ATTR-CM in different age groups were calculated and their 95% CI were estimated. For all tested hypotheses, the significance level of 0.05 was considered and multiple testing was adjusted by the Bonferroni correction method. All analyses were performed on R version 4.2.2.

## Results

Among the overall ATTR population reviewed (n=1802), 85 carriers were confirmed to have P/LP TTR variants without evidence of CM at the time of genetic testing. Specifically, 80 patients had normal echocardiography and 5 patients had negative PYP scintigraphy alone, 29 patients had both negative echocardiography and PYP scintigraphy. In addition, cMRI at baseline was normal in 4 patients.

### Baseline characteristics of TTR genotype-positive phenotype-negative individuals

Baseline characteristics of the patient population are reported in **Table 1**. The mean age was 48.5±11.7 years, 39% were males, and 91% were White. Comorbidities included spinal stenosis (2%), chronic kidney disease (1%), peripheral neuropathy (18%), and bilateral carpal tunnel syndrome (15%). One patient had undergone permanent pacemaker implantation and 2% had a prior history of syncope. Troponin levels were elevated in 17% of patients, and median NT-proBNP in the cohort was normal at 51 pg/ml (IQR 25-88). ECG abnormalities included atrioventricular block (n=3), atrial fibrillation (n=3), low voltage (n=1) and pseudo-infarct pattern (n=3). The median AI-enabled ECG probability of CA was 6.3% (IQR 1.9-19.3), and five patients had an AI-ECG probability above the threshold.

**Table 1.** Baseline characteristics of the overall G+/P-ATTRv cohort.

|  | Overall<br>(n=85) |
| --- | --- |
| Age (years) | 48.5 (11.7) |
| Male sex n (%) | 33 (39) |
| White n (%) | 77 (91) |
| Proband`s cardiac involvement n (%) | 47 (55) |
| Proband`s neurological involvement n (%) | 51 (60) |
| Proband`s organ involvement unknown n (%) | 10 (12) |
| First-degree relatives, n (%) | 63 (74) |
| Second-degree relatives, n (%) | 22 (26) |
| **Blood test** |  |
| WBC ($\times 10^3/\mu\text{L}$)<sup>a</sup> | 6.1 (5.1-6.9) |
| Hb (g/dL) | 13.9 (1.4) |
| PLT ($\times 10^3/\mu\text{L}$)<sup>a</sup> | 231.0 (208.0-251.5) |
| Positive troponin n (%) | 12 (17) |
| NTproBNP (pg/ml)<sup>a</sup> | 51.0 (25.0-88.0) |
| **Comorbidities** |  |
| Spinal stenosis n (%) | 2 (2) |
| Peripheral neuropathy n (%) | 15 (18) |
| CKD n (%) | 1 (1) |
| Bilateral carpal tunnel syndrome n (%) | 13 (15) |
| Pacemaker n (%) | 1 (1) |
| Syncope n (%) | 2 (2) |
| **ECG** |  |
| Atrial fibrillation n (%) | 3 (4) |
| AVB n (%) | 3 (4) |
| Low voltage n (%) | 1 (1) |
| Pseudo-infarct pattern n (%) | 3 (4) |
| AI amyloid probability above threshold n (%) | 5 (6) |
| **Imaging** |  |
| Echocardiography n (%) | 80 (94) |
| PYP scintigraphy n (%) | 34 (40) |
| Cardiac Magnetic Resonance n (%) | 4 (5) |
| **Baseline Echocardiographic Features** |  |
| LVEF | 62.6 (7.4) |
| Posterior wall thickness (mm) | 9.1 (1.4) |
| Septal wall thickness (mm) | 9.3 (1.5) |
| LVEDV (ml) | 127 (25.7) |
| LA dilation n (%) | 7 (10) |
| Biatrial dilation n (%) | 2 (3) |
| LAVI (ml/m<sup>2</sup>) | 28 (8.5) |
| RV dysfunction n (%) | 2 (3) |
| RVSP (mmHg) | 27.5 (5.2) |
| E/e`* | 7.7 (2.7) |
| LV GLS (%) | -20.5 (2.6) |
| **Gene variants** |  |
| Val30Met n (%) | 25 (29) |
| Thr60Ala n (%) | 24 (28) |
| Val122Ile n (%) | 8 (9) |
| Ile107Val n (%) | 6 (7) |
| Other n (%) | 22 (26) |

AI: artificial intelligence. AVB: atrioventricular block. CKD: chronic kidney disease. GLS: left ventricular global longitudinal strain. LVEDV: left ventricle end-diastolic volume. LVEF: left ventricle ejection fraction. LA: left atrium. LAVI: left atrium volume index. PYP: pyrophosphates. RV: right ventricle. RVSP: right ventricle systolic pressure.
\*: We reported the mean between septal and lateral E/e` when both were available. If lateral E/e` was not available, the only septal E/e` was reported.
Data represent mean (standard deviation, SD) or number (percentage) unless otherwise specified.
**a:** Data represent median (1<sup>st</sup> quantile, 3<sup>rd</sup> quantile).

None of the patients met the pre-defined echocardiographic criteria for ATTR-CM. Mean septal and posterior wall thickness were 9.3±1.5 and 9.1±1.4 mm respectively with a mean LV end-diastolic volume of 127 ml and LV ejection fraction (LVEF) of 62.6±7.4%. A sensitive indicator of systolic dysfunction, LV global longitudinal strain was slightly reduced (range -15% to -17%) in 11% of patients (n=7/61) but overall mean values were normal at -20.5±2.6%. Left atrial dilation was reported in 10% of subjects with a mean left atrium volume index that was normal at 28±8.5 ml/m2. Mean LV filling pressures were not significantly elevated (7.7 ± 2.7). The *TTR* P/LP variants observed in our population are listed in **Table S1** with Val30Met being the most prevalent (30%), followed by Thr60Ala (28%), Val122Ile (9%), and Ile107Val (7%).

### Clinical progression in TTR genotype-positive phenotype-negative carriers

Among the 85 G+/P-carriers, follow-up clinical and imaging studies were available in 55 patients. Clinical progression in this cohort is described in **Table 2**. Over a median follow-up period of 6.8 years (IQR 4.1 – 9.7), the risk of developing peripheral neuropathy during follow-up was 25.5% (95%CI: 8.9, 42.1) as compared to baseline (p=0.004). Specifically, there were an additional 4 patients who developed spinal stenosis, 7 bilateral carpal tunnel syndrome, 5 atrial fibrillation, and 6 syncope during follow-up as compared to baseline. Among the patients (n=24) who were diagnosed with peripheral neuropathy, 15 underwent skin/nerve biopsy, and 2 patients stained positive for amyloid deposition. Small interfering RNA or antisense oligonucleotide therapy was initiated in 12 patients.

**Table 2.** Clinical and echocardiographic characteristics in G+/P-ATTRv carriers who had follow-up data available (n=55).

|  | Baseline | Follow-up | Difference | p-value# |
| --- | --- | --- | --- | --- |
| **Comorbidities** |  |  |  |  |
| Spinal stenosis n (%) | 2 (3.6) | 6 (10.9) | 7.3 (-2.3, 16.9) | 0.142 |
| Peripheral neuropathy n (%) | 10 (18.2) | 24 (43.6) | 25.5 (8.9, 42.1) | 0.004 |
| CKD n (%) | 1 (1.8) | 6 (10.9) | 9.1 (0.1, 18.1) | 0.051 |
| Bilateral carpal tunnel syndrome n (%) | 9 (16.4) | 16 (29.1) | 12.7 (-2.8, 28.2) | 0.111 |
| Atrial fibrillation n (%) | 3 (5.5) | 8 (14.5) | 9.1 (-2, 20.2) | 0.112 |
| Pacemaker n (%) | 1 (1.8) | 1 (1.8) | - | - |
| Syncope n (%) | 2 (3.6) | 8 (14.5) | 10.9 (0.4, 21.5) | 0.047 |
| **Echocardiography** |  |  |  |  |
| LVEF | 63.3 (4.3) | 61.2 (3.6) | -2 (-3.6, -0.4) | 0.014 |
| Septal wall thickness (mm) | 9.4 (1.6) | 10.2 (2.4) | 0.7 (0, 1.3) | 0.037 |
| Abnormal septal wall thickness n (%) | 0 (0) | 4 (8.7) | 8.7 (0.6, 16.8) | 0.035 |
| Posterior wall thickness (mm) | 9.3 (1.5) | 9.2 (2.2) | 0 (-0.5, 0.6) | 0.853 |
| Abnormal posterior wall thickness n (%) | 0 (0) | 2 (4.5) | 4.5 (-1.6, 10.7) | 0.128 |
| Left atrium volume index (ml/m<sup>2</sup>) | 28.5 (9.6) | 29.2 (6.8) | 1.1 (-2.3, 4.5) | 0.518 |
| Left atrium dilation n (%) | 5 (10.2) | 14 (29.8) | 19.6 (4, 35.2) | 0.016 |
| Biatrial dilation n (%) | 2 (4.3) | 4 (8.9) | 4.6 (-5.5, 14.8) | 0.368 |
| E/e`* | 7.4 (2.2) | 7.2 (2.4) | 0.3 (-0.6, 1.1) | 0.558 |
| LV GLS (%) | -19.8 (2.7) | -20.4 (3) | -0.6 (-1.5, 0.3) | 0.205 |

CKD: chronic kidney disease. GLS: left ventricular global longitudinal strain. LV: left ventricular. LVEF: left ventricle ejection fraction.
\*: We reported the mean between septal and lateral E/e` when both were available. If lateral E/e` was not available, the only septal E/e` was reported.
Data represent mean (standard deviation, SD) or number (percentage) unless otherwise specified.
The proportional difference (95% confidence interval, CI) for binary variables or the mean difference (95% CI) for continuous variables are reported in the column named by "Difference".
For binary variables, the proportional difference between follow up and baseline levels was tested if it is different from 0. For continuous variables, paired t test was used to test if the variable's mean is different between baseline and follow up time. Tow-sided tests were used.

### Cardiac disease penetrance in TTR genotype-positive phenotype-negative carriers

Within this group, 53 underwent echocardiography, and 2 patients had a PYP scan alone, 18 had both PYP scintigraphy and echo, and 4 had both cMRI and echo at baseline. During follow-up, 12 (22%) of the 55 patients developed ATTRv-CM. Specifically, 5 patients developed a positive PYP scan, 6 patients were diagnosed based on the change in predefined echocardiography criteria and 1 patient developed cMRI features of CA. The disease-free at 10 years was estimated by the Kaplan-Meier method, and the 10-year risk of developing overt ATTRv-CM was 29.5% (95% CI: 7.9, 46.0, **Figure 1**). Additionally, cardiac penetrance was assessed among different age groups (**Table 3**). The penetrance increases as age increases. For the groups ≤ 50 years, 50-60 years, 60-70 years, and > 70 years, there are 11% (95% CI: 3%, 33%), 13% (95% CI: 4%, 38%), 33% (95% CI: 15%, 58%), and 43% (95% CI: 16%, 75%) patients developed ATTRv-CM at the end of the follow-up period, respectively. We also studied cardiac penetrance among different genetic variants. The age distribution is similar among the three most prevalent variants. The median ages at the end of follow-up are 61.0, 60.5, and 59.0 for patients who have Val130Met, Thr60Ala, and Val122Ile variants, respectively. The cardiac penetrance for the three genetic variants are 7% for Val130Met, 33% for Thr60Ala, and 40% for Val122Ile (**Table S2**).

**Figure 1.**
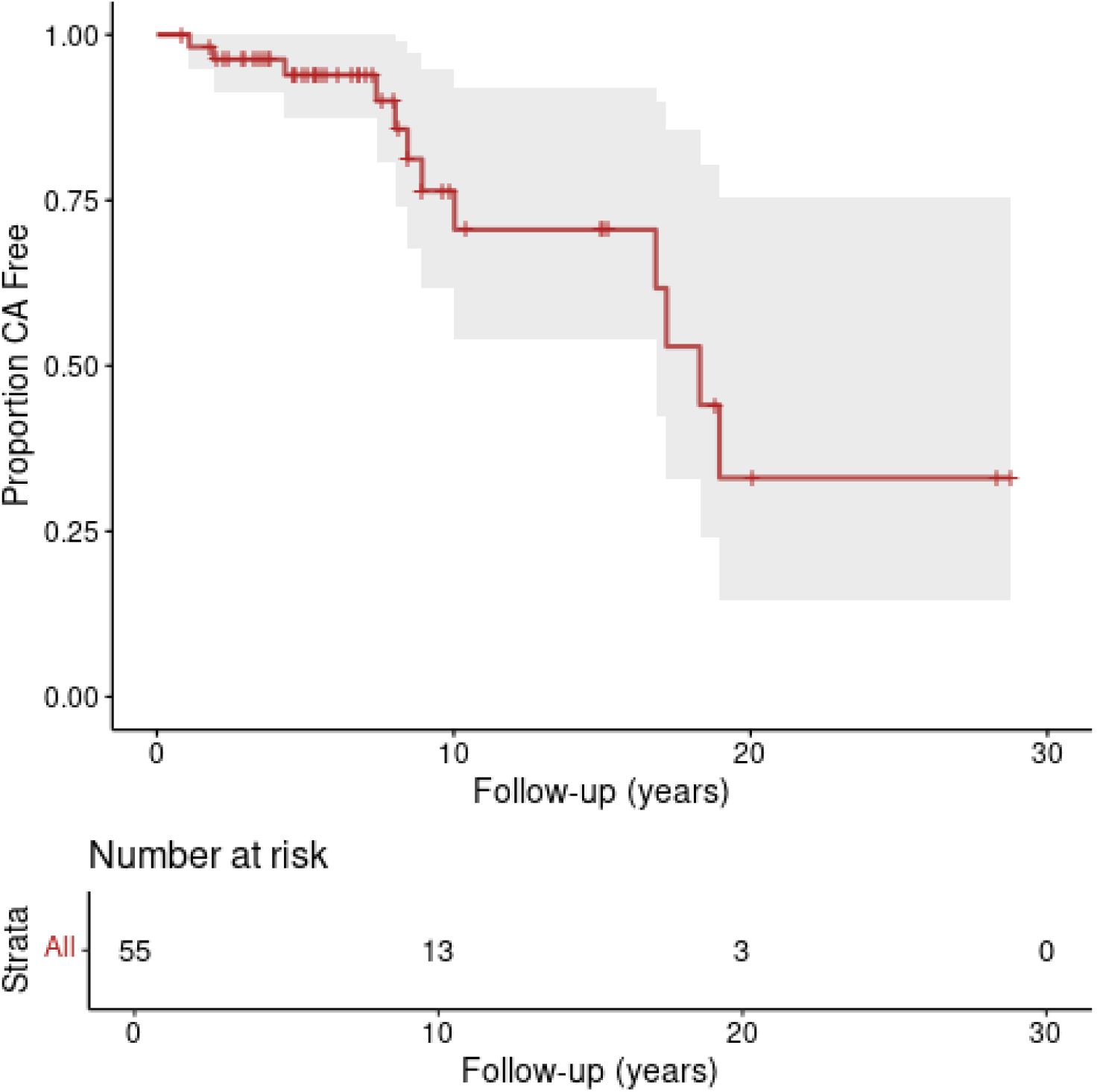
Kaplan-Meier curve showing the 10-year risk of developing overt ATTRv-CM (29.5%, 95% CI: 7.9, 46.0).

**Table 3.** Estimated penetrance for developing cardiac amyloidosis based on various G+/P-ATTRv carrier age groups.

| Age Groups* | Total No. of Patients | Penetrance | 95% CI |
| --- | --- | --- | --- |
| <= 50 years | 18 | 0.11 | (0.03, 0.33) |
| 50-60 years | 15 | 0.13 | (0.04, 0.38) |
| 60-70 years | 15 | 0.33 | (0.15, 0.58) |
| >= 70 years | 7 | 0.43 | (0.16, 0.75) |

\*: the age at the cardiac amyloidosis diagnosis or at the last follow-up.

### Echocardiographic changes in TTR genotype-positive phenotype-negative carriers

Echocardiographic progression in this cohort is described in **Table 2**. Overall, we observed a decrease in LVEF (p=0.014), an increase in septal wall thickness (p=0.037), and an increase in left atrium dilation proportion (p=0.016). Posterior wall thickness, left atrium volume index, bilateral dilation, E/è, and LV GLS did not significantly change. Notably, while at baseline no patients had abnormal wall thickness, 4 patients had abnormal septal wall thickness and 2 patients had abnormal posterior wall thickness during follow-up.

In the 12 patients diagnosed with ATTRv-CM (**Table 4**), the mean LVEF remained normal (60.3± 5.3%) but was reduced as compared to baseline (p=0.026). There was an increase in the mean septal wall (9.4 ± 1.0 mm to 12.0 ± 3.3 mm, p=0.019) and mean posterior wall thickness (9.8 ±1.4 mm to 10.9±3.3 mm, p=0.147). There was a significant increase in the proportion of ATTRv-CM patients who developed left atrium dilation at follow-up compared to baseline (63.6% vs 9.1%, p=0.008). Notably, 3 developed biatrial dilation during follow-up (p=0.038). Among the 43 patients who did not develop ATTRv-CM by pre-specified criteria (**Table S3**), there were expectedly no differences between any of the echocardiographic parameters during follow-up as compared to baseline.

**Table 4.** Differences between echocardiographic characteristics at baseline and last follow-up among G+/P-ATTRv carriers who developed CA (n = 12).

|  | Baseline | At CA diagnosis | Difference | p-value# |
| --- | --- | --- | --- | --- |
| LVEF | 65.2 (3.3) | 60.3 (5.3) | -5.2 (-9.6, -0.8) | 0.026 |
| Septal wall thickness (mm) | 9.4 (1) | 12 (3.3) | 2.7 (0.6, 4.8) | 0.019 |
| Abnormal septal wall thickness n (%) | 0 (0) | 4 (36.4) | 36.4 (7.9, 64.8) | 0.034 |
| Posterior wall thickness (mm) | 9.8 (1.4) | 10.9 (3.3) | 1.4 (-0.6, 3.4) | 0.147 |
| Abnormal posterior wall thickness n (%) | 0 (0) | 2 (22.2) | 22.2 (-4.9, 49.4) | 0.099 |
| Left atrium volume index (ml/m2) | 28.2 (10.6) | 33.3 (8.1) | 6.3 (-3.5, 16.1) | 0.173 |
| Left atrium dilation n (%) | 1 (9.1) | 7 (63.6) | 54.5 (21.4, 87.7) | 0.008 |
| Biatrial dilation n (%) | 0 (0) | 3 (33.3) | 33.3 (2.5, 64.1) | 0.038 |
| E/e`* | 8.1 (3.6) | 8.6 (4) | 2.2 (-1.4, 5.8) | 0.198 |
| LV GLS (%) | -19.3 (3.2) | -18 (4.1) | 0 (-3.4, 3.5) | 0.966 |

GLS: left ventricular global longitudinal strain. LV: left ventricular. LVEF: left ventricle ejection fraction.
\*: We reported the mean between septal and lateral E/e` when both were available. If lateral E/e` was not available, the only septal E/e` was reported.
Data represent mean (standard deviation, SD) or number (percentage) unless otherwise specified.
The proportional difference (95% confidence interval, CI) for binary variables or the mean difference (95% CI) for continuous variables are reported in the column named by "Difference".
For binary variables, the proportional difference between follow up and baseline levels was tested if it is different from 0. For continuous variables, paired t test was used to test if the variable's mean is different between baseline and follow up time. Tow-sided tests were used.

### Predictors of cardiac disease penetrance

Baseline characteristics were selected to evaluate for an association with ATTRv-CM development using Firth’s penalized logistic regression (**Table S4**). The proband’s age at the time of diagnosis (p-value = 0.0076) and AI-ECG amyloid probability (p-value = 0.0041) were associated with the development of ATTRv-CM. Next, a composite score was calculated that combined all the selected baseline characteristics to evaluate its predictive value for ATTRv-CM development. A Ridge penalized logistic regression was fitted using the disease development as a dependent variable and those baseline characteristics as independent variables. We use the estimated predictor of the fitted model as the composite score. We use the precision-recall analysis to evaluate the prediction performance of the composite score. The estimated AUC under the precision-recall curve from repeated cross-validation was 0.65, which indicated a prediction effect of those variables for ATTRv-CM development.

By fitting Cox proportional-hazard regressions, patient’s age (HR: 1.09, 95%: 1.01, 1.17, p=0.0246), proband’s age of diagnosis (HR: 0.91, 95%: 0.86, 0.98, p=0.0096), NTproBNP (HR: 1.00, 95%: 1.00, 1.01, p=0.0119), scaled AI ECG probability of ATTRv-CM (HR: 1.04, 95% CI: 1.01-1.07, p=0.0091), proband’s neurological involvement (HR: 0.23, 95% CI: 0.07-0.83, p=0.0244), and NTproBNP (HR: 1.01, 95% CI: 1.00-1.01, p=0.0187) were associated with the time to ATTR-CM development (**Table 5**). However, after adjusting for multiple testing these associations were not statistically significant.

**Table 5.** Association between baseline characteristics and ATTRv-CM development risk.

|  | Estimated HR | 95% CI | p-value# |
| --- | --- | --- | --- |
| Age (years) | 1.09 | (1.01,1.17) | 0.0246 |
| Male sex | 2.20 | (0.66, 7.33) | 0.1999 |
| Proband's age | 0.91 | (0.86, 0.98) | 0.0096 |
| Proband's cardiac involvement | 2.56 | (0.74, 8.80) | 0.1356 |
| Proband's neuro involvement | 0.23 | (0.07,0.83) | 0.0244 |
| NTproBNP (pg/ml) | 1.01 | (1.00,1.01) | 0.0187 |
| Peripheral neuropathy | 0.89 | (0.24, 3.36) | 0.8674 |
| Bilateral carpal tunnel syndrome | 3.78 | (0.89, 15.99) | 0.0705 |
| AI amyloid probability at ECG | 1.04 | (1.01, 1.07) | 0.0091 |
| LVEF | 1.01 | (0.89, 1.15) | 0.8778 |
| Posterior wall thickness (mm) | 1.41 | (0.86, 2.30) | 0.1736 |
| Septal wall thickness (mm) | 1.08 | (0.70, 1.66) | 0.7393 |
| Val30Met | 0.20 | (0.02, 1.56) | 0.1230 |
| Thr60Ala | 1.68 | (0.54, 5.23) | 0.3742 |
| Val122Ile | 4.96 | (0.91, 27.15) | 0.0648 |

AI: artificial intelligent; ECG: electrocardiogram; LVEF: left ventricle ejection fraction.
#: The raw p-value obtained from statistical tests are reported. Bonferroni correction was used to account for multiple testing. Therefore, the significance level is 0.05, and the adjusted significance level is 0.05/15 = 0.003.

## Discussion

This study, the largest of its kind to describe ATTRv G+/P-carriers, also sheds light on the complex dynamics of cardiac disease penetrance and expressivity in ATTRv, that could help counsel these individuals and may guide informed decision-making regarding the timing of initiating medical therapy. The observed cardiac penetrance rate of 22% over a median 6.8-year follow-up period and its age-dependency, underscores the latency and variable nature of disease manifestation among *TTR* variant carriers lacking structural evidence of CM at baseline. This study also provides insight into the evolution of cardiac morphology and function as assessed by echocardiography in this genetically diverse group of TTR carriers. Moreover, we observed that the proband’s age of diagnosis impacts on disease penetrance in their family members, the older the age at diagnosis of the proband, the lower the probability of developing amyloidosis in family members. We also generated the hypothesis of the potential use of AI-enabled ECG screening at baseline as a non-invasive method for predicting ATTRv-CM penetrance. The AI ECG screening tool offers a promising approach to risk stratification, perhaps enabling a precision medicine approach to attenuate disease progression and improve patient outcomes. Larger studies are needed to validate our exploratory observations.

Among genetic cardiomyopathies, the time from being diagnosed with a P/LP to developing the disease is highly variable and often undefined.^21–24^ A Large dataset demonstrated that the frequency of a P/LP *TTR* variant is not so rare (approximately 1 in 1000 individuals).^25^ The penetrance of ATTRv manifesting specifically as peripheral neuropathy is also variable in different geographical regions and among families.^26–28^ Similarly, ATTRv peripheral neuropathy is also characterized by incomplete cardiac penetrance and variable expressivity.^4^ A relationship between age at diagnosis and disease penetrance has been described in this disease, although the underlying age-dependent mechanisms are unknown.^29^ Male sex has also been associated with an increased risk for peripheral neuropathy disease penetrance in ATTRV30M carriers.^26^

The natural history of *TTR* variant carriers has been previously described in results of an observational survey but was based on occurrence of symptoms alone.^30^ The outcomes of “asymptomatic” patients (defined as absence of heart failure symptoms) with echocardiographic evidence of ATTR-CM at baseline have also been previously described,^31^ and individuals carriers of a P/LP *TTR* variant enrolled in the United Kingdom Biobank were found to be at higher risk of heart failure.^25^ However, information regarding ATTRv cardiac disease penetrance and expressivity based on cardiac imaging in G+/P-family members is largely unknown.^32^ In the Atherosclerosis Risk in Communities study, V122I *TTR* variant Black carriers (n=124) were found to be at an increased risk of incident heart failure as compared to non-carriers over 21.5 years of follow-up.^33^ Recently, V122I carriers who were predominately Black were described to have a higher risk of all-cause mortality and heart failure hospitalizations, particularly driven by heart failure with reduced EF, compared to non-carrier controls.^34^ However, in both these studies whether CA was present at baseline and whether heart failure occurred due to CA or other causes such as coronary artery disease or hypertension is unknown. Furthermore, White subjects were not studied given that the V122I variant is predominantly present in Blacks. Our study addresses these limitations by describing the disease-specific natural history of hereditary ATTR confirmed by imaging in G+P-patients who mostly had variants other than V122I.

It is relevant to highlight that 18% of our overall cohort of family members had a history of clinically diagnosed peripheral neuropathy at baseline and the nerve/skin biopsy, when performed, was most likely to be negative for amyloid deposition. The lack of sensitivity of the nerve/skin biopsies for amyloid maybe due to the multifocal amyloid deposition process, the early stage of the disease, and the possibility of peripheral neuropathy due to related comorbidities (three patients with negative biopsy had concomitant diabetes mellitus).^35,36^ However, the high prevalence and progression of peripheral neuropathy underscores the potential systemic nature of ATTRv amyloidosis and the importance of considering extracardiac manifestations in the risk assessment of these individuals. Despite we were not powered to predict the development of cardiac disease based on extracardiac manifestation, the presence of these extracardiac clinical manifestations should always raise the suspicion of systemic amyloidosis.

Increased LV wall thickness, a classic echocardiographic feature of CA, and a reduction in LVEF were observed but the latter variable remained within normal limits in G+/P-family members LVEF. Sensitive echocardiographic deformation parameters such as myocardial strain have been proposed to detect early involvement of the heart in amyloidosis. However, these values remained within the normal range in our population. Among the seven patients with abnormal global longitudinal strain at baseline, ATTRv-CM was diagnosed only in one case during follow-up. Therefore, global longitudinal strain may not be indicative of subsequent cardiac involvement in our population. Myocardial deformation echocardiographic imaging has primarily been validated in cohorts with some grade of increased myocardial wall thickness and whether this echocardiographic parameter could be useful in the early identification of ATTR-CM in a G+/P-population prior to this study is unknown.^37^ Moreover, we found that the age at which the proband is diagnosed with ATTRv influences cardiac penetrance in family members. This age-dependent penetrance finding in the proband aligns with the recommendation of beginning disease assessment in family members, 10 years before the affected proband’s age of disease onset.^10,38^

It is unknown whether and when to initiate newer therapy for ATTR amyloidosis in G+P-patients representing an unmet need in clinical practice. Our findings suggest that G+P-individuals with a positive AI-ECG and with an affected proband who was diagnosed at a younger age have a higher probability of developing CM. Patients with high AI-ECG probability may warrant more frequent follow-up with multimodality imaging, and cardiac biopsy may be indicated if imaging is inconclusive especially if patients are symptomatic to consider initiation of therapy.

### Patient’s perspective

Returning genetic testing results can be emotionally and logistically challenging for both family members and healthcare providers given the uncertainty in outcomes. Genetic testing results might have a profound impact on individuals and families, highlighting the need for such studies and high-touch methods such as one-on-one genetic counseling. We hope our study with others will help inform not only the clinicians who will encounter these patients but also the patients themselves.

### Limitations

A limitation of our study includes a relatively small sample size which is an inherent limitation of rare diseases, although it is the largest study of *TTR* P/LP variant carriers with longitudinal cardiac imaging follow-up that provides information on cardiac penetrance of ATTRv in the absence of clinically discernible cardiac disease. Although a longer period of follow-up may have yielded a higher cardiac disease penetrance, this study with a median 6.8-year follow-up period provides a reasonable estimate of disease risk. Furthermore, less than half had PYP scintigraphy, largely driven by the period of the study, which is known to be more sensitive than echocardiograms. Perhaps the cardiac phenotype penetrance might be even higher than what we observed. Larger cohorts will be required to validate our findings such as the use of AI-ECG to identify those at risk for ATTRv-CM development to enable early disease detection and intervention.

### Conclusion

In conclusion, the 10-year estimated risk of ATTRv-CM development in G+P-family members is high at 29.5%, and the most common echocardiographic features that characterize CM development are increased LV wall thickness and left atrial dilation. The risk of developing CM in ATTRv family members appears to be proband age-of-disease dependent and could be predicted by baseline AI ECG. These findings describe the high incidence of CA and the possibility of risk stratifying ATTRv G+P-patients not only to assist in counseling family members but also to provide guidance to clinicians regarding the timing of initiating appropriate but expensive therapy for ATTRv-CM. Further validation of our exploratory analysis results in larger cohorts is needed.

## Data Availability

Availability of all data is available via a request to the authors.

## NON-STANDARD ABBREVIATONS AND ACRONYMS

AI: artificial intelligence AI
ATTR: transthyretin amyloidosis
AUC: area under the curve
CA: cardiac amyloidosis
CI: confidence interval
CM: cardiomyopathy
CV: cross-validation
cMRI: cardiac magnetic resonance
ECG: electrocardiographic
HR: hazards ratio
LV: left ventricular
PR: precision recall
PYP: pyrophosphate

